# Characterizing Use of the Internet and Social Media to Access Suicide-Related Content among Adolescents with Suicidal Ideation

**DOI:** 10.64898/2026.10.13.26365422

**Authors:** Meredith Gansner, Prakruthi Nikam, Margaret Adams, Sai Ramrajesh, Nyah Huntley, William Wang, Mustafa Quadri, Zheala Qayuum

## Abstract

**Background:** Adolescents frequently consult online resources regarding sensitive health topics, including suicide. Although content analyses demonstrate that online platforms contain both suicide-preventive and pro-suicide material, they rarely incorporate subjective data from adolescents at elevated suicide risk.

**Aims:** This exploratory study examined associations between reasons for seeking suicide-related information online and the digital media used to access such content among adolescents with suicidal ideation.

**Method:** Over one year, 131 adolescents aged 13 to 18 with current or past suicidal ideation were recruited from the Emergency Department of an urban pediatric hospital to complete a cross-sectional survey. Participants reported whether they had sought suicide-related information online, the type of information sought, and the online locations used. Mixed-effects regression models examined associations between reasons for online searches and digital media used.

**Results:** Of 131 enrolled adolescents, 113 completed the survey; 69 (61.1%) reported ever seeking suicide-related information online. Searches were more often intended to assist with acting on suicidal thoughts than to reduce suicidal thoughts (63.7% vs. 55.1%). Social media were used for help-seeking searches slightly more often than professional health organization websites (42.1% vs. 31.6%). Among adolescents who used social media for their searches, discussion forums were significantly more likely to be used when seeking information to act on suicidal thoughts (OR=7.0, p=.02).

**Conclusion:** The majority of adolescents with suicidal ideation may seek out suicide-related content online, particularly to facilitate suicide planning. Social media may be used as frequently as professional mental health websites like 988 or The Trevor Project for suicide-related searches.

## Introduction

### Online Health Information Seeking and its Risks

It is estimated that about 9 out of 10 teenagers and young adults look online for health-related information, with approximately 40% searching for topics like anxiety or depression.(1) Mental health-related Internet searches are even more prevalent among youth with existing psychiatric illness,(1,2) and many adolescents use the Internet to explore and discuss high-risk behaviors, like drug use and suicide.(3) One study of young adults in the United Kingdom found that approximately half (48%) of those with a history of suicidal self-harm had ever looked online for information about suicide.(4)

Suicide and self-harm-related online searches are not without potential risk. Self-injurious behaviors can be “transmissible” through media, including digital platforms.(5–11) Engagement with non-suicidal self-injury online content can reinforce self-injurious behaviors(12–14) and at least one study has demonstrated suicide contagion via social media in a localized suicide cluster.(15) Furthermore, content analyses of suicide-related social media content warn that many posts do not adhere to the responsible suicide-reporting guidelines outlined by the American Federation of Suicide Prevention.(16,17)

### Addressing the Gaps

A 2025 screen of 97,000 U.S. teenagers found that a record 51% had experienced recent suicidal ideation,(18) and a growing number of adolescents may be searching online for information about suicide. However, few studies directly assess suicide-related online searches within populations of youth struggling with psychiatric illness. To best guide clinical care of suicidal adolescents, research is needed to determine types of suicide-related information most frequently sought in these online searches, as well as what online locations are being visited to access suicide-related content.

This pilot study examines suicide-related Internet searches among a sample of teenagers with current or history of suicidal ideation to identify: 1) what suicide-related information is being searched for online in this population of youth and 2) what online locations (e.g., discussion forums, mental health websites) are visited to access different types of suicide-related information. Based on prior studies examining digital locations where youth look for health-related information (both overall and mental health),(19,20) we expected websites and social media to be the platforms used most frequently to access suicide-related topics. Because content analyses also show a high-proportion of suicide-promoting content on social media,(17,21,22) we anticipated participants would preferentially use mainstream social media platforms to identify suicide methodology or solicit help in suicide planning.

## Methods

From 2024 to 2025, 131 youth were recruited from the Emergency Room of a tertiary pediatric hospital in an urban metropolitan region in the northeastern United States, where they had presented for emergent psychiatric evaluation. The authors assert that all procedures contributing to this work comply with the ethical standards of the relevant national and institutional committees on human experimentation and with the Helsinki Declaration of 1975, as revised in 2024. All procedures involving human subjects were approved by the Boston Children’s Research Institutional Review Board (approval number IRB-P00046908).

### Recruitment

#### Inclusion Criteria

Eligible participants were youth between the ages of 13 and 18 (including 18) who presented to the study site’s emergency department in psychiatric crisis and endorsed a lifetime history of suicidal ideation. History of suicidal ideation was determined by an affirmative answer to the first question of the Columbia Suicide Severity Rating Scale (C-SSRS) (i.e., “Have you ever wished you were dead or wished you could go to sleep and not wake up?”); the C-SSRS was administered to every patient undergoing a psychiatric assessment in the emergency department. Additionally, participants were only recruited if they were both eligible and at the study site between the hours of 8am and 6pm on a weekday (i.e., Monday through Friday) when study team members were on site.

#### Exclusion Criteria

Participants were not eligible for the study if they had a diagnosis of moderate or severe intellectual disability or active psychosis with a disorganized thought process. Participants were also not approached by the study team if they were exhibiting ongoing behavioral dysregulation (e.g., frequent shouting), psychomotor agitation (e.g., continuous pacing), or had required a restraint (either chemical and/or mechanical) within the previous twenty-four hours due to unsafe behaviors.

#### Informed Consent Process

Eligible individuals were identified by mental health clinicians on the Emergency Psychiatric Services (EPS) team (i.e., social worker, nurse practitioner or physician), who would assess patients who presented in psychiatric crisis. After completing their initial assessments, EPS clinicians would provide the names of eligible potential participants to the study team. If a potential participant was a minor, informed consent from a parent/guardian was required prior to enrollment. Parental/guardian consent was given either through written documentation or verbal phone consent if a parent/guardian was unable to be physically present at the time of enrollment. Written assent was required and documented for all minor participants. For participants aged 18 years, written informed consent was obtained and documented.

### Collected Data

Primary outcome survey questions can be reviewed in the manuscript’s supplementary material. All participant demographics were collected from the participant through the survey instrument, except for age, which was obtained through the hospital’s electronic health record system.

#### History of Online Searches for Suicide-Related Content

Participants were asked if they had ever looked up information about suicide online. If answered in the affirmative, the survey asked more specifically whether 1) this information had ever been accessed to find ways to reduce suicidal thoughts and/or 2) this information had ever been accessed to find ways to act on their suicidal thoughts. Participants were then asked for the specific online locations where they had looked for suicide-related information for both types of content searches.

#### Information Utility (for Help-Related Searches)

If a participant reported looking online for ways to reduce their suicidal thoughts, they were also asked if they felt that the material they accessed did reduce their suicidal ideation (yes/no). If a participant reported having used the Internet for this purpose multiple times with differing results, they could enter both a “yes” and “no” response. Participants were also asked whether they had ever used the Internet to message/text suicide prevention resources (e.g. 988 Suicide and Crisis Lifeline or The Trevor Project) for help in not acting on their suicidal thoughts.

### Survey Administration and Safety Flag Protocol

Study team members administered the survey to all participants verbally and recorded each answer into the hospital’s secure REDCap system. Each participant represented an individual survey response. This method of administration was chosen for two reasons: 1) hospital patients in psychiatric crises were not permitted access to internet-enabled smart devices (e.g. an iPad) once a need for psychiatric hospitalization was determined and, 2) patients in the emergency room awaiting placement for transfer to an inpatient psychiatric facility were not permitted to have sharp tools like staples and paper clips, which would have been required using a traditional “pencil and paper” survey methodology.

Due to the potential mental health risks associated with the viewing of suicide-related content, participants were asked whether they had searched for suicide-related online content at any time during the week prior to their emergency room presentation. All affirmative responses were reported to the study PI directly following survey completion, who would communicate this finding to the responsible clinician on the participant’s hospital treatment team and document the positive “safety flag” in the participant’s hospital medical record. Because this information could influence treatment recommendations if previously undisclosed, positive safety flags were relayed to the participant’s treatment team within a few hours of survey completion. During the consent process, participants (and legal guardians when applicable) were informed that the survey contained safety flag questions where the research team would be required to alert the hospital team if a participant answered in the affirmative.

### Data Processing

Primary outcomes and predictors of interest were binarily coded with “1” indicating that the variable was present and “0” that the variable was absent. Online locations provided by participants were sorted into broader, pre-defined categories (e.g., social media and professional health organization websites like 988 or The Trevor Project).

Digital media categories were established based on prior research regarding adolescent use of the Internet for health information, metrics of presumed reliability of accessed information (professional health websites), presence or absence of social interaction (social networking sites), and community “membership” determined by a shared interest (social networking sites). Use of three specific social media subtypes was assessed: general blogging/networking (e.g., Facebook, X/Twitter), predominantly audiovisual media sharing (e.g., Instagram, TikTok), and discussion forums (e.g., Reddit, Quora). Popular mainstream media-sharing platforms have been demonstrated to feature images and videos with suicide and self-injury methods,(12,14,17) content that may perpetuate self-harm,(12,23) and in their systematic review of Internet use and self-harm, Marchant et al. summarizes the literature on self-harm discussion in online forums, citing concerns that individuals are encouraged to act on suicide plans and given detailed information about suicide methods.(23)

Participants were also given the opportunity to input a free-text answer into an additional “Other” category if they had sought suicide-related information through a digital media subtype that did not fall under the pre-established categories. For each digital media subtype a participant reported using, the participant was asked also to provide the name(s) of the specific platform (e.g., TikTok, Reddit).

### Data Analyses

Descriptive, demographic variables were calculated as percentages. Because some participants were expected to have performed online searches to both act on and reduce their suicidal thoughts, some observations would not be independent; therefore, multilevel logistic regression models were performed to examine relationships between digital media used for online searches and type of information sought. Analyses were performed using Stata 19.5.

## Results

There were 131 participants enrolled in the survey study and 113 (86.3%) completed surveys. The incomplete surveys were considered “missing at random” because they resulted from technological issues with data collection on the part of the study team, rather than participant non-response or withdrawal from the study. Survey participants were predominantly female sex (n=81, 72.3%) and white, non-Hispanic/Latine (n=57, 50.4%), with a mean age of 15.0 (SD=1.5) (**Table 1**).

**TABLE 1:**
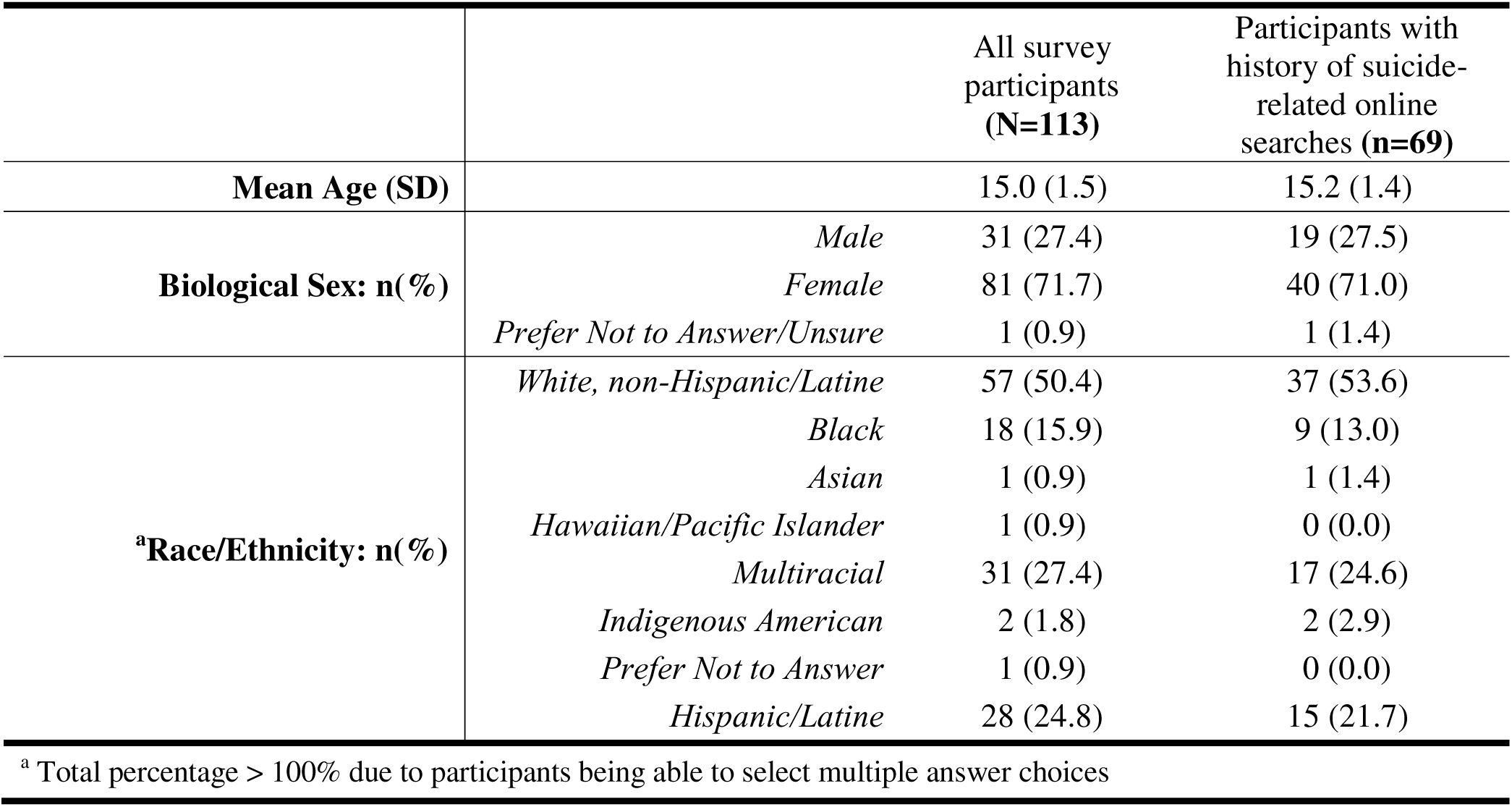
SAMPLE DEMOGRAPHICS.

### Frequency of Suicide-Related Online Searches and Digital Media Used

**Figure 1** shows the percentages of participants who reported ever looking up suicide-related content online, and for what purpose(s). Out of the 113 participants with completed surveys, 69 (61.1%) reported ever having sought suicide-related content online. These searches were made most frequently to identify information to act on suicidal thoughts (n=44, 63.8%). Nearly one-third of participants with a history of suicide-related online searches reported seeking both information to act on and to reduce suicidal thoughts (n=20/69, 29.0%).

**Figure 1:**
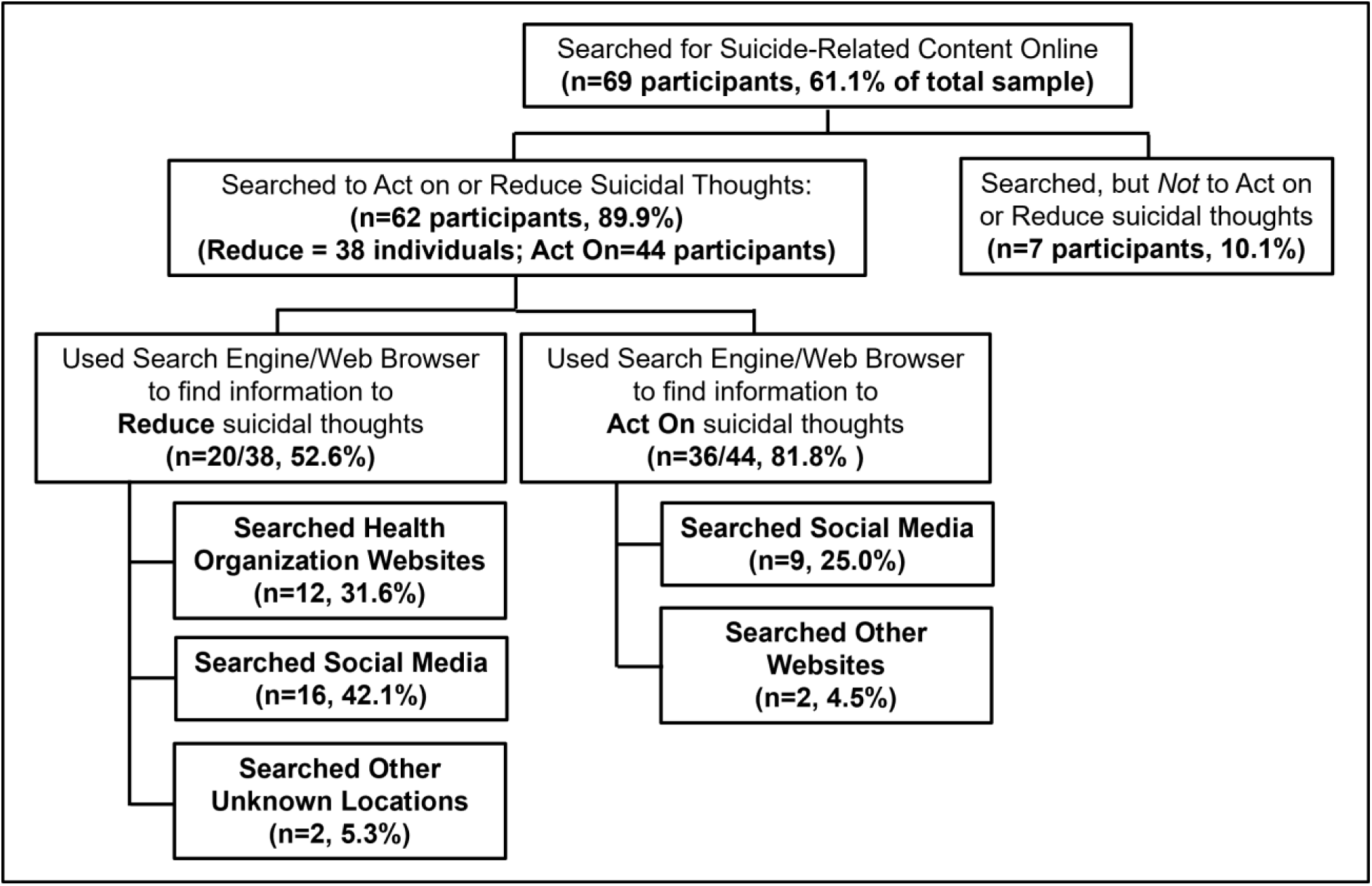
Schematic depicting proportion of sample who engaged in online searches for suicide-related content, including categories of digital media used for searches. Participants were instructed to list all digital media used for searches and to consider all such searches they had made previously.

Overall, 45 participants used an online search engine to start their query (e.g., Google Search): for help-seeking searches, 52.6% (n=20/38), and for searches to act on suicidal thoughts, 81.8% (n=36/44) (**Figure 1**). Social media were used by 16 participants to access online help for reducing suicidal thoughts (42.1% of participants who made such searches) and 17 participants when seeking information to act on suicidal thoughts (38.6% of participants who made such searches). Websites affiliated with established suicide-prevention organizations or healthcare organizations (regional or national) were accessed by 12 of the 38 (31.6%) participants who had searched online for help with reducing suicidal thoughts. Half of those 12 participants (n=6) reported accessing the 988 Suicide and Crisis Lifeline website specifically, and two of those teenagers (2/6, 33.3%) also cited use of Google to perform a suicide-related online search. In **Figure 2**, the specific online locations and social media that were used to access suicide-related information are delineated. Online forums were the most frequently used type of social media to look up suicide methodology (n=14/44, 31.8%), while audiovisual media-sharing social media platforms were the most frequently used type of social media to seek help for reducing suicidal thoughts (n=10/38, 26.3%).

**Figure 2:**
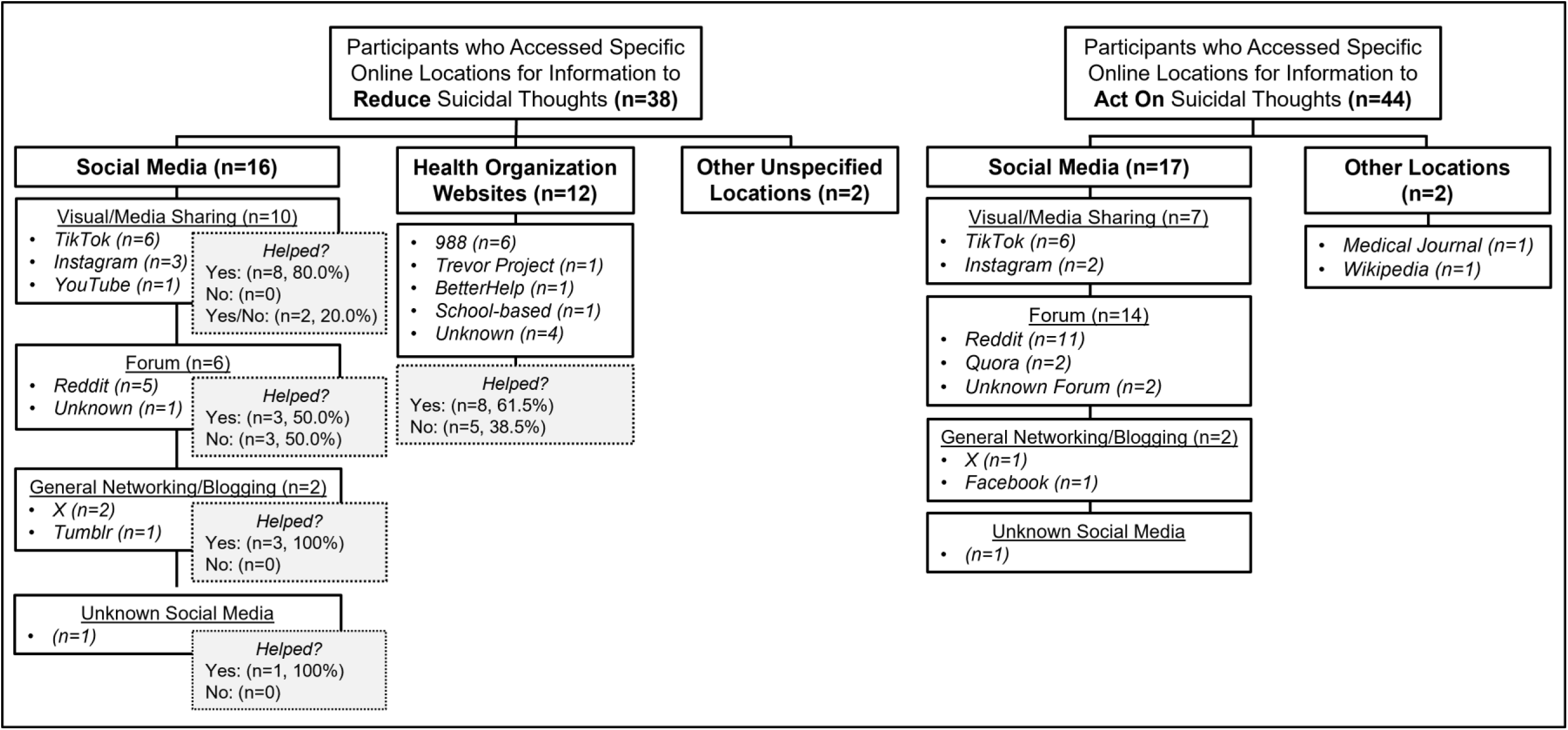
Schematic depicting online locations accessed in searches of suicide-related content to reduce vs. act on suicidal thoughts. Participants were instructed to list all digital media used for searches and to consider all such searches they had made previously.

Of the 69 adolescents who reported searching for suicide-related content online, only 2 (2.9%) used digital media to message a counselor/trained professional at any mental health crisis line; a single individual reported texting or messaging the 988 Suicide and Crisis Lifeline specifically.

### Digital Media Used for Online Searches Based on Type of Information Sought

In online searches seeking suicide methods, participants were significantly more likely to report use of a search engine in their query (OR: 4.05, CI: 1.50-11.0, p=*0.006) (**Table 2**). Social media were no more likely to be used over other digital media when searching to act on vs. reduce suicidal thoughts. However, comparing use of social media subtypes among only those participants who used social media in their searches, discussion forums were significantly more likely to be accessed when searching for information to act on suicidal thoughts (OR: 7.0, CI: 1.39-35.3, p=0.02) (**Table 2**).

**TABLE 2:** Digital Media Associated with Online Searches to Act on vs. Reduce Suicidal Thoughts.

|  | % of participants<br>who used in searches<br>to <u>ACT ON</u> suicidal<br>thoughts | % of participants<br>who used in searches<br>to <u>REDUCE</u> suicidal<br>thoughts |  |  |  |  |
| --- | --- | --- | --- | --- | --- | --- |
| SEARCH ENGINE<br>(vs. no search engine used) | n (%) | n (%) | OR | CI | SE | p<sup>f</sup> |
|  | 36/44 (81.8%) | 20/38 (52.6%) | 4.05 | 1.50-11.0 | 2.06 | *0.00 |
| ANY SOCIAL MEDIA<br>(vs. other online locations) | n (%) | n (%) | OR | CI | SE | p<sup>f</sup> |
|  | 17/44 (38.6%) | 16/38 (42.1%) | 0.87 | 0.36-2.10 | 0.39 | 0.75 |
| SOCIAL MEDIA TYPE vs.<br>OTHER ONLINE LOCATIONS | n (%) | n (%) | OR | CI | SE | p<sup>f</sup> |
| *General/Blogging<sup>a,d</sup>* | 2/43 (4.65%) | 2/37 (5.40%) | 0.85 | 0.45-3.25 | 0.88 | 0.88 |
| *Media-Sharing<sup>b,d</sup>* | 7/43 (61.3%) | 10/37 (38.7%) | 0.53 | 0.18-1.56 | 0.29 | 0.25 |
| *Discussion Forum<sup>c,e</sup>* | 14/44 (31.8%) | 6/37 (16.2%) | 2.41 | 0.82-7.10 | 1.33 | 0.11 |
| COMPARING SOCIAL MEDIA<br>TYPES ONLY | n (%) | n (%) | OR | CI | SE | p<sup>f</sup> |
| *General/Blogging vs. Other Social Media<sup>d</sup>* | 2/16 (12.5%) | 2/15 (13.3%) | 0.93 | 0.11-7.59 | 1.00 | 0.95 |
| *Media-Sharing vs. Other Social Media<sup>d</sup>* | 7/16 (43.8%) | 10/15 (66.7%) | 0.39 | 0.09-1.67 | 0.29 | 0.21 |
| *Discussion Forum vs. Other Social Media<sup>e</sup>* | 14/17 (82.4%) | 6/15 (40.0%) | 7.0 | 1.39-35.3 | 5.78 | *0.02 |

<sup>a</sup> Facebook, Twitter/X, Tumblr
<sup>b</sup> TikTok, YouTube, Instagram
<sup>c</sup> Quora, Reddit
<sup>d</sup> Participant who reported use of one unspecified social media platform in both searches to act on & reduce suicidal thoughts and use of a forum *only* in searches to act on suicidal thoughts excluded from analyses
<sup>e</sup> Omitted participant included in count/analysis of participants who used forums in searches to act on suicidal thoughts
<sup>f</sup> \* Denotes significance with p < 0.05

## Discussion

Existing research has demonstrated both the ease with which a teenager can access suicide-related content online as well as prevalence of suicide-related content with pro-suicide messaging.(17,22) However, it is unknown how many teenagers with lifetime suicidal ideation seek out this type of online content, where the information is sought, and for what purpose(s). Our results suggest that the Internet is an important source of suicide-related information for adolescents with significant mental health concerns. Our results highlight some concerning findings: in youth who have experienced suicidal ideation, 1) suicide-related online content may be accessed more for suicide planning than for seeking help, 2) reflexive mental health supports (e.g., 988 contact information) generated automatically by search engines may not prompt help-seeking behavior, and 3) online forums may be preferentially visited for the purpose of suicide planning.

### Prevalence of Suicide-Related Online Searches

Few studies examine prevalence of suicide-related Internet searches among samples of youth with suicidal ideation, particularly in the U.S. One 2008 study of Japanese youth found that only 15.9% (33/208) of participants with a history of suicidal ideation had accessed information about suicide online.(24) A more recent 2015 UK study found that 48.0% of young adults who had made prior suicide attempts reported having ever made a suicide-related Internet search.(4) Because the number of adolescents and young adults who use digital media to acquire new information has increased over time,(25,26) this likely explains our increased prevalence of 61.1%. It is likely that little (if any) suicide-related research occurs offline in this age group.

There is even less existing research on what suicide-related information is most sought by youth during these searches. Thus, while multiple studies provide evidence of youth using the Internet to access both help-seeking and suicide planning content,(23) we have no data with which to compare our proportions of adolescents with prior suicidal ideation who had searched to act on thoughts or to reduce them. However, the percentage of participants in our sample who endorsed online suicide planning specifically (38.9%) is similar to at least one prior estimate of the proportion of youth with lifetime suicidal ideation participate in suicide planning (33.4%).(27)

### Use of Social Media vs. Professional Websites for Help-Seeking Searches

While necessary to interpret in the context of both limited geographic scope and sample size, our results suggest that youth with suicidal ideation may visit social media more than professional health organization websites to find help for their suicidal thoughts. If similarly confirmed in other regions, this is consistent with growing evidence that youth are preferentially turning to social media over more accredited, and potentially, reliable online sources.(19) Surveys have found that 81% of 13-17-year olds use social media influencers as their source of news and information,(28) and at least one research team has demonstrated this tendency when youth search online for information about high-risk behavior. In their 2012 study, Duggan et al. found peer-created sites appear to be accessed more frequently for information about non-suicidal self-injury than sites created by mental health professionals.(29)

In our sample, fewer than a third of participants who had ever sought help for suicidal thoughts online did so via professionally-created websites, a finding that could be the result of several different factors. For one, adolescents in this high-risk group could not be aware of, or familiar with, professionally-created mental health sites, although this seems improbable given that both regional and national mental health resources were highly visible in this study’s geographical region, particularly to youth with psychiatric diagnoses. Alternatively, these adolescents may be looking for help that they do not believe is available through professionally-created resources. For example, adolescents may be seeking information from others with disclosed lived experience. And, no less relevant, via the process of assortative relating, youth with psychiatric illness often form relationships with others who have similar psychological struggles.(11) It is possible that some youth avoid using professional resources as a source of help for their suicidal thoughts because of peers’ negative experiences with these platforms, such as perceived inefficacy or breach in confidentiality if personal information was disclosed due to safety concerns.

### Use of the 988 Website and Text Messaging Option

Similarly, while our survey did not ask about calls to the 988 Suicide and Crisis Lifeline, few participants listed the 988 website as a source of help or reported ever using text message to access 988. Data collected following launch of the 988 Lifeline showed that adolescents were the most-represented age group among the Lifeline’s text message and online chat users,(30) but our results suggest that the proportion of in-need adolescents who actually connect to 988 may still be relatively small.

Also arguing against the hypothesis that study participants may not have been aware of professional resources is the overwhelming percentage of youth who reported using Google in their search for suicide-related information. There were 45 respondents in total who reported using Google search to find suicide-related content (45/46, 97.8%), and the single participant who listed only a web browser for their online search used Safari, which defaults to Google as its primary search engine.

Because Google automatically responds to suicide-related queries with the statement “Help is available” and links to the 988 website and hotline, it initially appeared encouraging that Google was a common “starting point” for adolescents’ suicide-related online searches. Over half of the study participants (n=46) who reported searching for suicide-related content online had been shown alternatives to suicide, regardless of the reason for their Internet search. However, still only six of those adolescents (6/46, 13.0%) accessed the 988 Lifeline website, and only one contacted 988 via text or chat message (2/46, 2.2%).

### Variability of Digital Media Used for Online Help-Seeking vs. Online Suicide-Planning

Correlations between the type of digital media used and type of suicide-related Internet search highlight how different platform capabilities address the differing goals of the adolescent user. For example, when our study’s participants looked online for ways to reduce their suicidal thoughts, they were significantly less likely to use online forums and search engines than if they were looking to identify suicide methods. Online forums and search engines do not necessarily offer direct messaging in real-time, unlike TikTok and Instagram, the social media platforms used most by our respondents. Whether consciously or unconsciously, help-seeking participants avoided looking for content on platforms that had less opportunity for interpersonal connection.

Alternatively, when searching for suicide methods, users may gravitate towards optimizing information quantity and relevance. Online search engines offer instant access to a wide variety of sources focused on a particular topic, although many of the results may adhere to AFSP guidelines. Online forums, on the other hand, give users access to active threads and thread archives containing granular details about a specific topic, including high-risk topics like drug purchasing and suicide. Content is moderated inconsistently across forums, with more permissive platforms becoming online environments where controversial viewpoints can be discussed openly and appear normalized through the creation of “echo chambers.”(31) Online forums have previously been linked to accessible suicide-related content,(4,22) as well as suicide planning in cases of suicide deaths.(17–19) Notably, a 2011 study examining where youth are most likely to see suicide stories online found online discussion forums to be the only source associated with greater suicidal ideation in the viewer;(20) it may also be that discussion forums are more commonly sought out for suicide-related content when youth are experiencing more significant suicidal thoughts.

As researchers study the relative impacts of suicide-related content exposure across different online platforms, it will also be important to clarify whether the odds of using a specific platform increase with the severity of mental health symptoms. For example, if the phenomenon of suicide contagion appears more pronounced on online forums, it may be that individuals selectively spend more time on online forums when they’re increasingly thinking about suicide planning.

### Limitations

As mentioned previously, the results of this study need to be considered in the context of several limitations, the most significant of which are the single site nature of the study and small sample size. While the study setting could impact generalizability of our findings, our sample’s demographics share many characteristics with recent national statistics on pediatric emergency room visits for psychiatric crises:(32) predominantly female gender, 50.4% identifying as white non-Hispanic (vs. 44% among U.S. boarding nationally for psychiatric crises). Furthermore, trends in adolescent social media use do not appear to change significantly based on geographic area, suggesting that teenagers with suicidal ideation elsewhere in the United States have access to the same types of digital content.(33)

The small sample size limits our ability to examine relationships between suicide-related content searches and demographic characteristics in greater detail (e.g., analyses comparing participants with no history of suicide-related online searches to those who only sought suicide-related online content for suicide-planning purposes). It may also increase the degree to which our findings were influenced by common limitations associated with self-report survey studies (e.g., social desirability and recall bias). In addition to surveying a larger number of participants, future studies should consider 1) recruiting youth from sites where survey answers might be less likely to influence treatment decisions surrounding psychiatric hospitalization (i.e., psychiatrically hospitalized adolescents) and 2) focusing on suicide-related searches within a specific timeframe (e.g. days before a suicide attempt).

## Conclusion

As the amount of information available online continues to grow, so will the proportion of adolescents who use the Internet to look up content related to high-risk behaviors, such as suicide. However, it is less certain to what extent adolescents will be able to appraise content veracity or recognize how the structure of online platforms can influence individual behavior. Our results support the need for additional research examining prevalence of suicide-related online searches as well as choice (or rejection) of specific online platforms when seeking help in managing suicidal ideation. Future studies that examine the temporal relationship between online searches for suicide methodology and youth suicide attempts are also critical; resulting data could inform both measures to reduce youth access to the most high-risk digital content as well as improve the way youth are directed to mental health support online.

## Acknowledgements

We would like to thank Dr. Micah Breiger for his help with recruitment. Artificial Intelligence use: No artificial intelligence tools have been used in the research or writing process for this manuscript.

## Data Availability Statement

The data that support the findings of this study are available from the corresponding author upon reasonable request.

## Author contributions

**MG, PN, SR, NH, WW, MQ** contributed to data acquisition and project design. **MG, MA and ZQ** contributed to project design, data analyses, and data interpretation. All authors contributed to the drafting of the manuscript and approved the final version.

## References

1. Rideout V, Fox S. Digital Health Practices, Social Media Use, and Mental Well-Being Among Teens and Young Adults in the U.S. Well Being Trust; 2018. p. 96. (Articles, Abstracts, and Reports). Report no.: 1093.

2. Chang FC, Chang C, Tao CC. Adolescent pursuit of health information online during the COVID-19 pandemic: The roles played by eHealth literacy and psychological distress. Camb Prisms Glob Ment Health. 2023;10:e51. doi:10.1017/gmh.2023.44

3. Carew C, Kutcher S, Wei Y, McLuckie A. Using Digital and Social Media Metrics to Develop Mental Health Approaches for Youth. Adolesc Psychiatry. 2014 Jul 31;4(2):116–21. doi:10.2174/221067660402140709122825

4. Mars B, Heron J, Biddle L, Donovan JL, Holley R, Piper M, et al. Exposure to, and searching for, information about suicide and self-harm on the Internet: Prevalence and predictors in a population based cohort of young adults. J Affect Disord. 2015 Oct;185:239–45. doi:10.1016/j.jad.2015.06.001

5. Niederkrotenthaler T, Stack S, Till B, Sinyor M, Pirkis J, Garcia D, et al. Association of Increased Youth Suicides in the United States With the Release of *13 Reasons Why*. JAMA Psychiatry. 2019 Sep 1;76(9):933. doi:10.1001/jamapsychiatry.2019.0922

6. Jack B. Goethe’s Werther and its effects. Lancet Psychiatry. 2014 Jun;1(1):18–9. doi:10.1016/S2215-0366(14)70229-9

7. Randall JR, Nickel NC, Colman I. Contagion from peer suicidal behavior in a representative sample of American adolescents. J Affect Disord. 2015 Nov;186:219–25. doi:10.1016/j.jad.2015.07.001

8. Bridge JA, Greenhouse JB, Ruch D, Stevens J, Ackerman J, Sheftall AH, et al. Association Between the Release of Netflix’s 13 Reasons Why and Suicide Rates in the United States: An Interrupted Time Series Analysis. J Am Acad Child Adolesc Psychiatry. 2020 Feb;59(2):236–43. doi:10.1016/j.jaac.2019.04.020

9. Gould MS, Kleinman MH, Lake AM, Forman J, Midle JB. Newspaper coverage of suicide and initiation of suicide clusters in teenagers in the USA, 1988–96: a retrospective, population-based, case-control study. Lancet Psychiatry. 2014 Jun;1(1):34–43. doi:10.1016/S2215-0366(14)70225-1

10. Gould M, Jamieson P, Romer D. Media Contagion and Suicide Among the Young. Am Behav Sci. 2003 May;46(9):1269–84. doi:10.1177/0002764202250670

11. Hawton K, Hill NTM, Gould M, John A, Lascelles K, Robinson J. Clustering of suicides in children and adolescents. Lancet Child Adolesc Health. 2020 Jan;4(1):58–67. doi:10.1016/S2352-4642(19)30335-9

12. Brown RC, Fischer T, Goldwich AD, Keller F, Young R, Plener PL. #cutting: Non-suicidal self-injury (NSSI) on Instagram. Psychol Med. 2018 Jan;48(2):337–46. doi:10.1017/S0033291717001751

13. Whitlock JL, Powers JL, Eckenrode J. The virtual cutting edge: The Internet and adolescent self-injury. Dev Psychol. 2006 May; Children, Adolescents, and the Internet 42(3):407–17. Located at: 2006-07128-003. doi:10.1037/0012-1649.42.3.407

14. Lewis SP, Heath NL, St Denis JM, Noble R. The Scope of Nonsuicidal Self-Injury on YouTube. Pediatrics. 2011 Mar 1;127(3):e552–7. doi:10.1542/peds.2010-2317

15. Swedo EA, Beauregard JL, de Fijter S, Werhan L, Norris K, Montgomery MP, et al. Associations Between Social Media and Suicidal Behaviors During a Youth Suicide Cluster in Ohio. J Adolesc Health. 2021 Feb;68(2):308–16. doi:10.1016/j.jadohealth.2020.05.049

16. Khasawneh A, Chalil Madathil K, Dixon E, Wiśniewski P, Zinzow H, Roth R. Examining the Self-Harm and Suicide Contagion Effects of the Blue Whale Challenge on YouTube and Twitter: Qualitative Study. JMIR Ment Health. 2020 Jun 9;7(6):e15973. doi:10.2196/15973

17. Molly Rose Foundation. Pervasive-by-Design: TikTok and Instagram [Internet]. Molly Rose Foundation; 2025. p. 1–40. Available from: https://mollyrosefoundation.org/wp-content/uploads/2025/08/proof3_PervasivebyDesign.pdf

18. Mental Health America. Youth reporting frequent thoughts of suicide reached record high in 2025, screening data show [Internet]. Alexandria, Virginia; 2026 [cited 2026 Sep 1]. Available from: https://mhanational.org/news/youth-reporting-frequent-thoughts-of-suicide-reached-record-high-in-2025-screening-data-show/

19. Stifjell K, Sandanger TM, Wien C. Exploring Online Health Information–Seeking Behavior Among Young Adults: Scoping Review. J Med Internet Res. 2025 Sep 9;27:e70379. doi:10.2196/70379

20. Dunlop SM, More E, Romer D. Where do youth learn about suicides on the Internet, and what influence does this have on suicidal ideation?: Influence of the Internet on suicidal ideation. J Child Psychol Psychiatry. 2011 Oct;52(10):1073–80. doi:10.1111/j.1469-7610.2011.02416.x

21. Picardo J, McKenzie SK, Collings S, Jenkin G. Suicide and self-harm content on Instagram: A systematic scoping review. Triberti S, editor. PLOS ONE. 2020 Sep 2;15(9):e0238603. doi:10.1371/journal.pone.0238603

22. Biddle L, Derges J, Mars B, Heron J, Donovan JL, Potokar J, et al. Suicide and the Internet: Changes in the accessibility of suicide-related information between 2007 and 2014. J Affect Disord. 2016 Jan;190:370–5. doi:10.1016/j.jad.2015.10.028

23. Marchant A, Hawton K, Stewart A, Montgomery P, Singaravelu V, Lloyd K, et al. A systematic review of the relationship between internet use, self-harm and suicidal behaviour in young people: The good, the bad and the unknown. PLOS One. 2017 Aug;12(8). doi:10.1371/journal.pone.0181722

24. Katsumata Y, Matsumoto T, Kitani M, Takeshima T. Electronic media use and suicidal ideation in Japanese adolescents. Psychiatry Clin Neurosci. 2008 Dec;62(6):744–6. doi:10.1111/j.1440-1819.2008.01880.x

25. Wartella E, Rideout V, Montague H, Beaudoin-Ryan L, Lauricella A. Teens, Health and Technology: A National Survey. Media Commun. 2016 Jun 16;4(3):13–23. doi:10.17645/mac.v4i3.515

26. Commonsense Media. News and America’s kids: How Young People Perceive and are Impacted by the News [Internet]. 2017. Available from: www.commonsensemedia.org

27. Nock MK, Green JG, Hwang I, McLaughlin KA, Sampson NA, Zaslavsky AM, et al. Prevalence, correlates and treatment of lifetime suicidal behavior among adolescents: Results from the National Comorbidity Survey Replication – Adolescent Supplement (NCS-A). JAMA Psychiatry. 2013 Mar;70(3). doi:10.1001/2013.jamapsychiatry.55 PubMed PMID: 23303463; PubMed Central PMCID: PMC3886236.

28. The Media Insight Project. The evolving news landscape: Comparing media habits and trust between teens and adults [Revenue & Resilience Study] [Internet]. 2026 Apr [cited 2026 Aug 31]. p. 1–53. Available from: https://americanpressinstitute.org/comparing-news-consumption-trust-us-teens-and-adults/

29. Duggan JM, Heath NL, Lewis SP, Baxter AL. An Examination of the Scope and Nature of Non-Suicidal Self-Injury Online Activities: Implications for School Mental Health Professionals. School Ment Health. 2012 Mar;4(1):56–67. doi:10.1007/s12310-011-9065-6

30. Stangland S. 988’s Texting, Chatting Options Give Youths in Crisis a Lifeline [Internet]. 2025 Jan [cited 2026 Feb 10]. (American Academy of Pediatrics News). Available from: https://publications.aap.org/aapnews/news/30952/988-s-texting-chatting-options-give-youths-in?autologincheck=redirected

31. Lerman K, Chu MD, Bickham C, Luceri L, Ferrara E. Safe spaces or toxic places? Content moderation and social dynamics of online eating disorder communities. Epj Data Sci. 2025;14(1):55. doi:10.1140/epjds/s13688-025-00575-5 PubMed PMID: 40726606; PubMed Central PMCID: PMC12296748.

32. Center for Disease Control and Prevention. Changes in Pediatric Emergency Department Visits Associated with Mental Health Conditions Before and During the COVID-19 Pandemic — United States, January 2019–January 2022. 2022 Feb. p. 1–12. (Weekly Report: Morbidity and Mortality Weekly Report (MMWR)).

33. Vogels EA, Gelles-Watnick R, Massarat N. Teens, Social Media and Technology 2022. Washington, D.C: Pew Research Center; 2022 Aug.

