## Supplementary Table 1 for "Characterizing Use of the Internet and Social Media to Access Suicide-Related Content among Adolescents with Suicidal Ideation"

**Have you ever looked up information related to suicide online?**

**Have you ever looked up information related to suicide online in order to find ways to reduce or NOT ACT on your suicidal thoughts?**

**WHERE ONLINE have you looked up information related to suicide in order to find ways to reduce or not act on your suicidal thoughts?**

**Did looking up suicide-related information on [PARTICIPANT-REPORTED ONLINE LOCATION] help you find ways to reduce or not act on your suicidal thoughts?**

**Have you ever looked up information related to suicide ONLINE in order to research how you might act on your suicidal thoughts or make a suicide plan?**

**WHERE ONLINE have you looked up information related to suicide in order to research how you might act on your suicidal thoughts or make a suicide plan?**

Supplementary Table 1: Wording of Survey Question for Primary Study Outcomes
